# Vascular geometry and network topology reveal vascular remodelling beyond density measures across Chew grades in macular telangiectasia type 2

**DOI:** 10.64898/2026.09.13.26361127

**Authors:** Michael Hafner, Daniel Deschler, Siegfried G. Priglinger, Maximilian J. Gerhardt

**Affiliations:** Department of Ophthalmology, LMU University Hospital, LMU Munich, Mathildenstrasse 8, 80336, Munich, Germany

**Keywords:** macular telangiectasia type 2, optical coherence tomography angiography, Chew classification, foveal avascular zone, vessel length density, capillary topology

## Abstract

**Background/Objectives:** Previous OCTA studies across MacTel severity levels have primarily characterised perfusion- and density-based vascular changes. Whether graph-derived network architecture provides complementary information on severity remains unclear. We therefore tested whether lower Chew grades already show architectural abnormalities despite comparatively preserved global vessel density.

**Subjects/Methods:** This cross-sectional study included 62 MacTel and 64 healthy control eyes. Fovea-centred 3×3-mm high-resolution swept-source OCTA acquired with the DREAM platform was analysed with OCTAVA. Whole-image vessel length density (VLD), regional vessel area density (VAD), peri-FAZ perfusion, foveal avascular zone (FAZ) geometry and network topology were assessed using mixed-effects models adjusted for age, sex and axial length.

**Results:** Grades 0-2 already showed greater FAZ acircularity than controls in the superficial (SCP; +0.132; q<0.001) and deep capillary plexus (DCP; +0.073; q=0.034), with lower SCP branchpoint density (q=0.022). Across MacTel grades, DCP VLD decreased by 0.352 mm^−1^ per grade (95% CI -0.583 to -0.121; Holm P=0.007), whereas the SCP slope was not significant (Holm P=0.071; plexus interaction P=0.160). DCP parafoveal VAD declined by 0.593 percentage points per grade (95% CI, -0.943 to -0.242; q=0.007), with no comparable decline in foveal VAD. Visual acuity worsened by 0.071 logMAR per grade (P<0.001).

**Conclusions:** OCTA suggests two complementary components of the MacTel vascular phenotype: geometric and topologic remodelling in lower grades, with additional regional and global capillary rarefaction in higher grades. Vascular geometry and network topology, therefore, provide complementary information not captured by vessel density alone.

## 1 Introduction

Macular telangiectasia type 2 (MacTel) is a bilateral neurodegenerative macular disease in which Müller-cell dysfunction, photoreceptor loss and metabolic disturbance coexist with characteristic parafoveal vascular abnormalities.[1–3] The historical Gass and Blodi stages describe visible vascular and pigmentary findings,[4] whereas the multi-modal Chew classification grades eyes from 0 to 6 using structural optical coherence tomography (OCT), fundus autofluorescence, colour imaging and neovascular features.[5–7] This scale provides a clinically interpretable framework for disease severity, but its quantitative retinal vascular correlates remain incompletely defined.

Optical coherence tomography angiography (OCTA) resolves the superficial capillary plexus (SCP) and deep capillary plexus (DCP) without dye leakage. Prior work has demonstrated telangiectatic dilation, capillary rarefaction, right-angled vessels, outer-retinal vascular extension and associations with ellipsoid-zone loss.[8–14]

Recent large-cohort work has characterised retinal and choriocapillaris density across the Chew classification, establishing quantitative perfusion associations with disease severity.[15, 16] Most recently, Stettler et al.[16] provided a comprehensive swept-source OCTA characterisation across the full Chew scale, demonstrating grade-associated changes in retinal vessel density and choriocapillaris flow deficits in a large cohort. However, whether Chew severity is also reflected in the organisation of the retinal capillary network itself, and whether such architectural abnormalities are detectable when global vessel density remains comparatively preserved, remains unclear. To our knowledge, graph-derived network topology, including branchpoint density, node connectivity and segment length, has not previously been examined across the Chew classification.

We therefore asked whether quantitative OCTA across Chew grades supports a density-only model of vascular severity or a broader phenotype in which vascular geometry and network architecture are already abnormal in lower grades. Using standardised high-resolution 3 *×* 3-mm swept-source OCTA and an open-source analysis pipeline, we quantified whole-image vessel length, regional and peri-FAZ perfusion, FAZ geometry and network topology. We hypothesised that lower grades would show geometric or topologic abnormalities despite comparatively preserved global VLD, whereas higher grades would show additional regional and global capillary rarefaction. Identifying an architectural signature that precedes measurable density loss would extend the window in which OCTA-derived biomarkers could inform disease monitoring or stratification for future interventional trials.

The DREAM OCTA platform used here has previously demonstrated detailed visualisation of the retinal microvasculature, particularly in the DCP, including detection of smaller-calibre vessels and complex branching patterns compared with several established OCTA systems.[17]

## 2 Subjects and methods

### 2.1 Study design and participants

This prospective cross-sectional observational study recruited participants at the Department of Ophthalmology, LMU University Hospital, Munich, Germany, from November 2024 through January 2025. The Institutional Review Board of the Faculty of Medicine, LMU Munich, approved the protocol (24-0571). The study adhered to the Declaration of Helsinki; all participants provided written informed consent; and the reporting follows the STROBE statement.[18]

MacTel was diagnosed by clinical examination and multimodal imaging. Healthy controls had no retinal or choroidal disease. Exclusion criteria were another retinal or choroidal disorder, diabetes mellitus, media opacity, insufficient signal, decentration, major motion artefact, or segmentation failure persisting after manual correction.

### 2.2 Imaging and disease grading

Each eye underwent a comprehensive ophthalmic examination and axial length measurement using the IOLMaster 700 (Carl Zeiss Meditec, Jena, Germany). Fovea-centred 3 *×* 3-mm OCTA volumes (512 *×* 512 pixels) were acquired with a 200-kHz swept-source DREAM OCT VG200D system (Intalight Inc., San Jose, CA, USA). Manufacturer-generated en face images were exported for the SCP and DCP using the device slab definitions.[19] Segmentation was visually inspected and corrected when necessary.

Two trained graders (M.H. and M.J.G.), masked to quantitative OCTA results, independently assigned Chew grades 0-6 from multi-modal images; discrepancies were resolved by consensus.[5]

### 2.3 OCTA image analysis

Images were analysed with the open-source OCTA Vascular Analyser (OCTAVA) in MATLAB R2024b (MathWorks, Natick, MA, USA), following Untracht et al.[20] and the device-specific workflow previously reported for DREAM OCTA.[17] A two-dimensional Frangi filter with a maximum kernel size of 3 pixels was followed by fuzzy-means thresholding; no median filter was applied. Binarised images were skeletonised, network connectivity was represented as an undirected graph, and isolated elements or branches shorter than a twig size of 2 pixels were removed. Automated FAZ masks and intermediate segmentations were visually reviewed.

Whole-image vessel length density (VLD; mm-1) was the principal density endpoint. Regional vessel area density (VAD; %) was quantified in the foveal circle, parafoveal rim, quadrants and superior/inferior hemi-rings. FD300 represented vessel density within a 300-*µ*m ring surrounding the FAZ. Network endpoints comprised branchpoint density, node count and mean segment length. FAZ endpoints included area, acircularity index and eccentricity; additional geometric variables were retained for supportive analyses.

### 2.4 Statistical analysis

Both eyes were retained. Linear mixed-effects models included a participant-level random intercept and fixed effects for age, sex and axial length. Within MacTel, Chew grade was modelled primarily as an ordinal continuous term. A grade-by-plexus interaction tested whether VLD slopes differed between the SCP and DCP. A clinically motivated transition contrast compared grades 4-6 with grades 0-3 because grade 4 introduces OCT hyperreflectivity in the Chew classification.[5]

For comparisons with healthy controls, MacTel eyes were grouped as early (grades 0-2), intermediate (3-4) and advanced (5-6), matching the graphical severity strata. Multiplicity was controlled using the Holm correction for paired primary/transition contrasts and the Benjamini-Hochberg false-discovery-rate correction for families of regional, FAZ, and topology endpoints. Sensitivity analyses excluded grade 6 and added logMAR visual acuity. A separate structure-function analysis examined visual acuity across Chew grade and whether DCP VLD or parafoveal VAD was independently associated with logMAR after adjustment for grade. Two-sided P<0.05 after the prespecified correction was considered significant. Analyses were performed in R version 4.4.2 (R Foundation for Statistical Computing, Vienna, Austria).

### 2.5 Generative artificial intelligence

OpenAI ChatGPT (OpenAI, San Francisco, CA, USA) assisted with statistical code review and English-language editing. It was not used to acquire or alter data, assign clinical grades or generate synthetic study images. The authors independently verified all analyses, source citations and manuscript content and accept full responsibility for the work.

## 3 Results

### 3.1 Cohort

The analysis included 126 eyes: 62 MacTel eyes from 31 participants and 64 healthy eyes from 32 controls (Table 1). Mean age was 62.3 (SD 10.5) years in MacTel and 60.0 (SD 11.1) years in controls; 18/31 (58.1%) and 21/32 (65.6%) participants, respectively, were female. Mean axial length was 22.30 (SD 1.09) mm in MacTel eyes and 23.53 (SD 0.96) mm in controls. Median visual acuity in MacTel was 0.25 logMAR (IQR 0.10-0.40). Chew-grade counts for grades 0-6 were 8, 5, 21, 2, 8, 13 and 5 eyes, respectively.

**Table 1.** Participant and eye characteristics.

| Characteristic | Healthy controls | MacTel |
| --- | --- | --- |
| Participants, n | 32 | 31 |
| Eyes, n | 64 | 62 |
| Age, years, mean (SD)* | 60.0 (11.1) | 62.3 (10.5) |
| Female participants, n (%) | 21 (65.6) | 18 (58.1) |
| Axial length, mm, mean (SD)† | 23.53 (0.96) | 22.30 (1.09) |
| Visual acuity, logMAR, median (IQR)† | 0.00 | 0.25 (0.10-0.40) |
| Chew grade 0, eyes, n (%) | — | 8 (12.9) |
| Chew grade 1, eyes, n (%) | — | 5 (8.1) |
| Chew grade 2, eyes, n (%) | — | 21 (33.9) |
| Chew grade 3, eyes, n (%) | — | 2 (3.2) |
| Chew grade 4, eyes, n (%) | — | 8 (12.9) |
| Chew grade 5, eyes, n (%) | — | 13 (21.0) |
| Chew grade 6, eyes, n (%) | — | 5 (8.1) |

IQR, interquartile range; SD, standard deviation. \*Participant-level summary. †Eye-level summary; inferential analyses accounted for within-participant correlation.

### 3.2 Whole-image capillary rarefaction across Chew grades

Whole-image VLD provided the global severity signal (Figure 1). Within MacTel, DCP VLD decreased by 0.352 mm^*−*1^ per grade (95% CI -0.583 to -0.121; Holm P=0.007). The SCP estimate was smaller and did not reach corrected significance (-0.222 mm^*−*1^ per grade; 95% CI -0.463 to 0.020; Holm P=0.071). The grade-by-plexus interaction was not significant (P=0.160), so the data do not support a formal difference in grade slopes between plexuses.

**Fig. 1.**
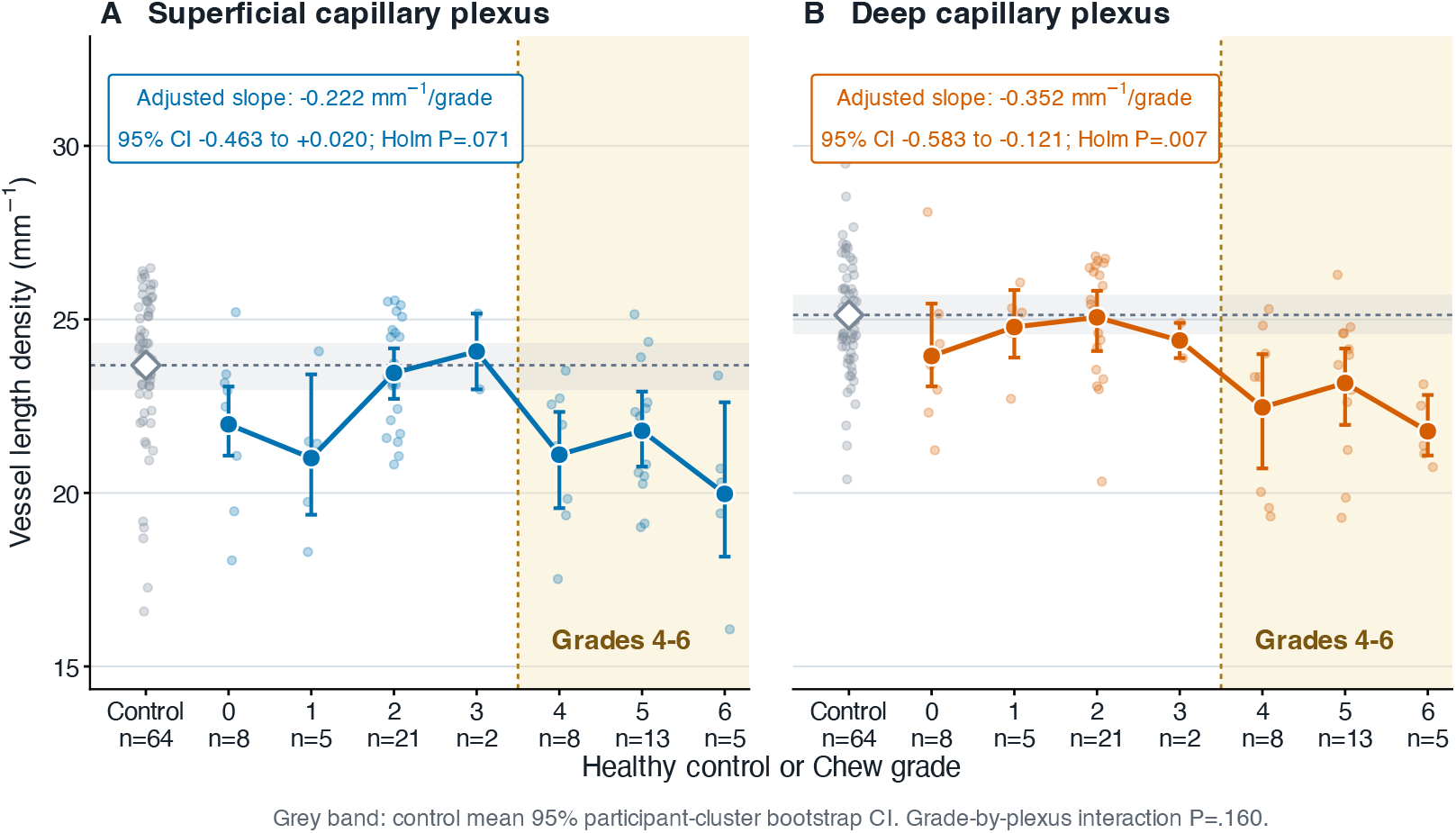
Whole-image vessel length density across Chew grades and the healthy-control reference. Eye-level VLD is shown for the superficial capillary plexus (A) and deep capillary plexus (B). Small points represent individual eyes; coloured points with error bars represent grade-specific means with participant-cluster bootstrap 95% confidence intervals. The grey dashed line and band show the healthy-control mean and 95% participant-cluster bootstrap confidence interval. The shaded region marks grades 4-6. In the report, covariate-adjusted ordinal mixed-model slopes are presented. The grade-by-plexus interaction was not significant (P=0.160).

Consistent with the ordinal DCP association, the prespecified grades 4-6 versus 0-3 contrast showed lower VLD in both plexuses in the standardised multi-domain analysis (Supplementary Figure S1). After excluding grade 6, the DCP transition remained evident (-1.171 mm^*−*1^; 95% CI -2.177 to -0.165; P=0.024), whereas the corresponding SCP estimate was imprecise (-0.801 mm^*−*1^; 95% CI -1.797 to 0.195; P=0.112).

### 3.3 Regional and peri-FAZ perfusion

Regional analyses showed that this higher-grade rarefaction was spatially non-uniform rather than a uniform fall in perfusion (Figure 2 and Supplementary Figure S2). DCP parafoveal VAD decreased by 0.593 percentage points per grade (95% CI -0.943 to -0.242; q=0.007), whereas foveal VAD showed no evidence of decline (+0.228; 95% CI -0.506 to 0.962; q=0.715). Accordingly, the parafoveal-minus-foveal DCP VAD slope was -0.945 percentage points per grade (95% CI -1.633 to -0.256; Holm P=0.016).

**Fig. 2.**
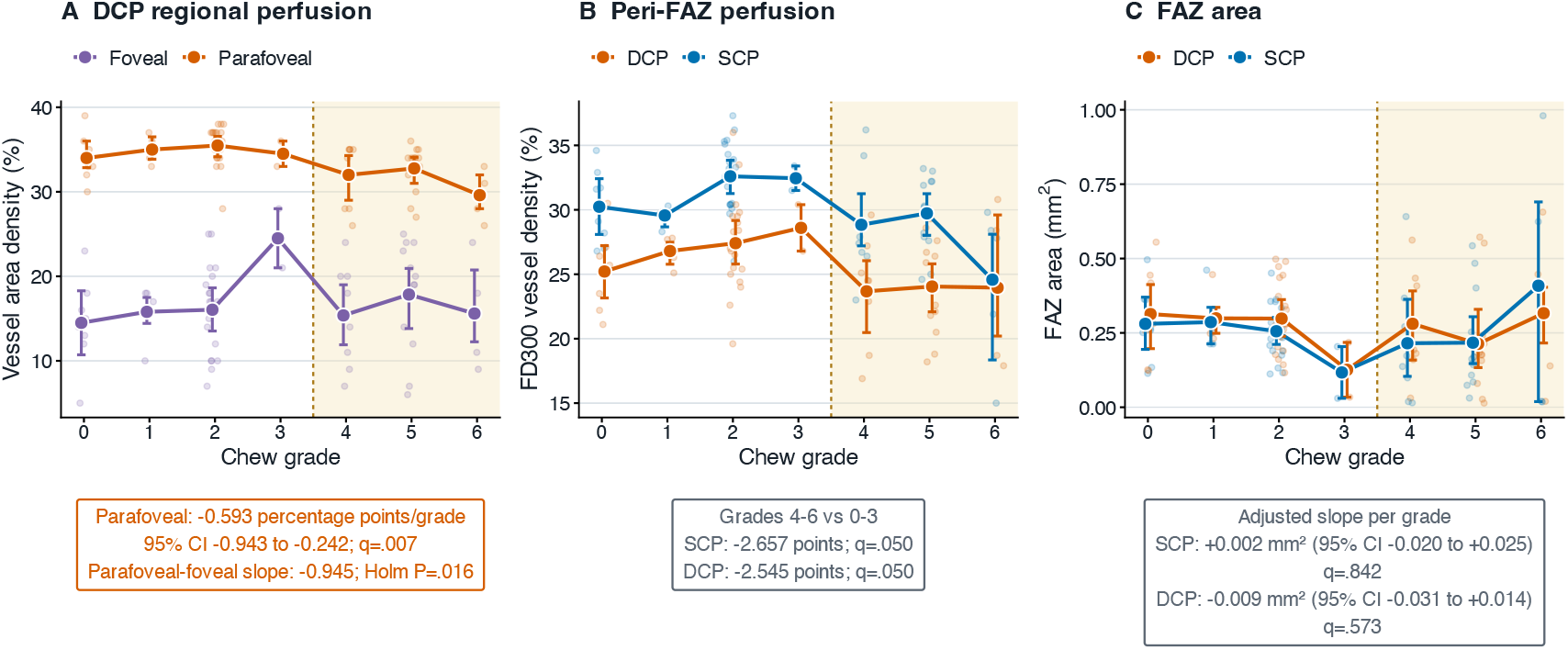
Regional and peri-FAZ perfusion across Chew grades. (A) Deep capillary plexus foveal and parafoveal vessel area density; the parafoveal slope was -0.593 percentage points per grade (95% CI -0.943 to -0.242; q=0.007), and the parafoveal-minus-foveal slope was -0.945 percentage points per grade (Holm P=0.016). (B) FD300 vessel density around the FAZ; grades 4-6 versus 0-3 were lower in the SCP (-2.657 percentage points; q=0.050) and DCP (-2.545; q=0.050). (C) The FAZ area showed no significant ordinal slope in either plexus. Points represent eyes, and larger symbols with error bars show grade-specific means with confidence intervals.

Among regional sectors, DCP VAD declined most consistently in the nasal quadrant (-0.813 percentage points per grade; 95% CI -1.233 to - 0.394; q=0.005), inferior hemi-ring (-0.715; 95% CI -1.133 to -0.298; q=0.007) and temporal quadrant (-0.584; 95% CI -1.031 to -0.137; q=0.037). The SCP showed a significant nasal decline (-0.674; 95% CI -1.141 to -0.206; q=0.022). Peri-FAZ FD300 was lower for grades 4-6 than 0-3 in both layers (DCP -2.545 percentage points, 95% CI -4.478 to -0.611; q=0.050; SCP -2.657, 95% CI -4.621 to - 0.693; q=0.050), while FAZ area itself showed no ordinal association in either layer.

### 3.4 Lower-grade geometric and network remodelling

The density findings did not capture the full lower-grade phenotype. FAZ shape abnormalities were already detectable in grades 0-2 (Figure 3): compared with controls, acircularity was higher in the SCP (+0.132; 95% CI 0.083 to 0.181; q<0.001) and DCP (+0.073; 95% CI 0.018 to 0.127; q=0.034). SCP eccentricity increased with Chew grade by 0.042 per grade (95% CI 0.022 to 0.062; q<0.001), whereas the DCP slope was not significant (+0.012; 95% CI -0.011 to 0.036; q=0.525).

**Fig. 3.**
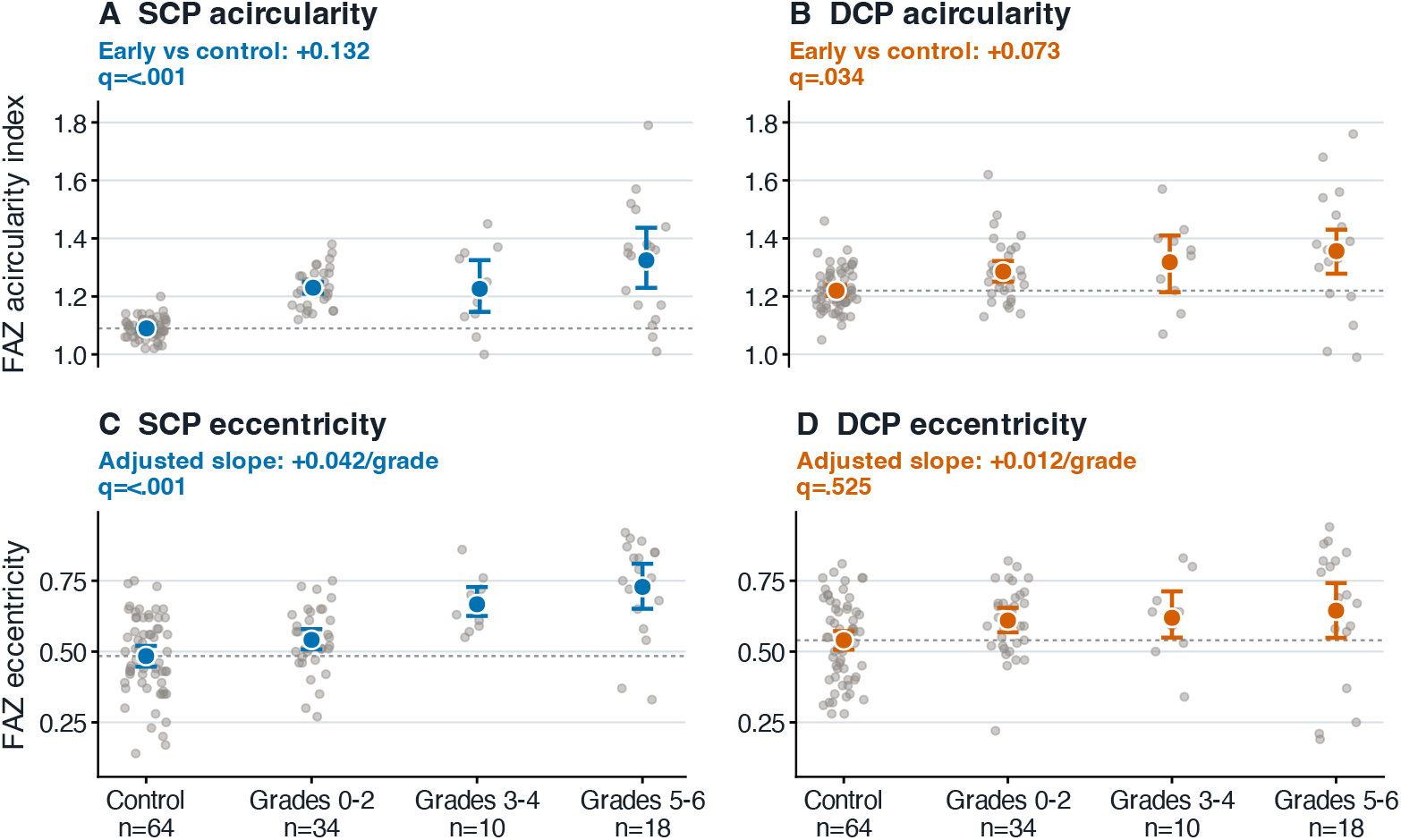
FAZ shape across healthy controls and MacTel severity strata. Acircularity index is shown for the SCP (A) and DCP (B), and eccentricity for the SCP (C) and DCP (D). Severity groups are early (Chew grades 0-2), intermediate (3-4) and advanced (5-6). Early grades already showed higher acircularity than controls in the SCP (+0.132; q<0.001) and DCP (+0.073; q=0.034). SCP eccentricity increased by 0.042 per Chew grade (q<0.001); the DCP slope was not significant (+0.012; q=0.525). Error bars are 95% confidence intervals from adjusted mixed-effects models.

Network topology provided an independent view of the same lower-grade remodelling (Figure 4). In grades 0-2, SCP branchpoint density was lower than in controls (-0.446; 95% CI -0.792 to -0.100; q=0.022), SCP node count was lower (-182.679; 95% CI -352.558 to -12.800; q=0.049), and mean SCP segment length was greater (+1.330; 95% CI 0.149 to 2.510; q=0.042). By grades 5-6, both plexuses showed lower branchpoint density and node counts with longer surviving segments (all q*≤*0.005). Thus, lower-grade eyes already differed in vascular shape and network organisation, whereas higher grades additionally showed broader network simplification and rarefaction.

**Fig. 4.**
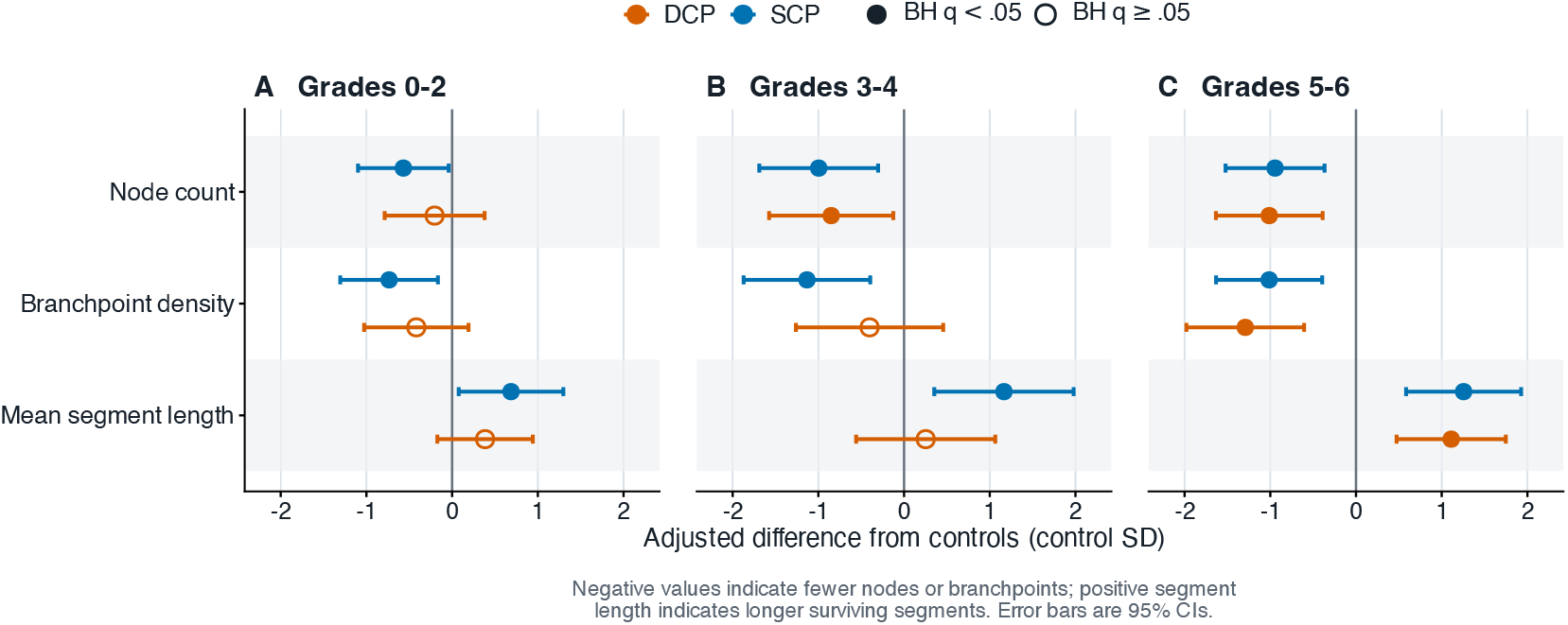
Network topology differences from healthy controls across MacTel severity strata. Adjusted differences for node count, branchpoint density and mean segment length are standardised to the control standard deviation separately for each endpoint. Filled symbols indicate Benjamini-Hochberg q<0.05 and open symbols q *≥* 0.05; error bars are 95% confidence intervals. Negative node and branchpoint estimates indicate network simplification, whereas positive mean segment length indicates longer surviving vascular segments. SCP, superficial capillary plexus; DCP, deep capillary plexus.

### 3.5 Visual function

Visual acuity worsened by 0.071 logMAR per Chew grade (95% CI 0.036 to 0.107; P<0.001). Adding visual acuity to the DCP VLD severity model preserved the VLD association (-0.339 mm^*−*1^ per grade; 95% CI -0.596 to -0.083; P=0.010). Conversely, neither DCP whole-image VLD nor DCP parafoveal VAD remained independently associated with logMAR after ordinal Chew grade was added (Holm P=0.797 for both). In this cross-sectional cohort, vascular and visual abnormalities therefore tracked the broader multimodal severity state more clearly than either single DCP metric tracked acuity independently.

## 4 Discussion

This study identifies two complementary components of the retinal vascular phenotype across the Chew classification. First, lower-grade MacTel was already distinguished from healthy eyes by FAZ deformation and superficial network simplification despite comparatively preserved whole-image vessel length. Second, higher grades were associated with additional regional and global capillary rarefaction, with the clearest corrected ordinal association in the DCP. The central implication is therefore not merely that vascular density decreases with severity, but that density, geometry and network architecture capture different aspects of the same disease phenotype.

The lower-grade phenotype is consistent with qualitative OCTA descriptions of temporal DCP abnormalities and perivenular capillary loss,[8, 12] and with multilayer studies showing vascular changes adjacent to outer-retinal abnormalities in early MacTel.[13] Our data add a quantitative architectural dimension: FAZ acircularity and superficial branchpoint/node metrics already differed from controls in grades 0-2. Global VLD can remain comparatively preserved when vessels are displaced, pruned, or regionally reorganised; this explains why shape and topology can reveal disease-related vascular remodelling that is not apparent from a single whole-field density measure.

The higher-grade phenotype was characterised by increasingly regional and global loss of perfused capillary signal. DCP parafoveal VAD declined across grades while foveal VAD did not, and the most consistent regional slopes involved the nasal quadrant and inferior hemi-ring, with a smaller temporal effect. Peri-FAZ FD300 was also lower across the grades 3-4 transition despite no corresponding ordinal change in FAZ area. Together, these findings argue against a spatially uniform reduction in perfusion and favour progressive involvement of specific parafoveal vascular compartments across increasing cross-sectional severity.

Stettler et al.[16] recently provided a comprehensive quantitative OCTA characterisation across Chew grades, demonstrating changes in retinal vessel density, FAZ metrics and choriocapillaris flow deficits. Our findings complement this work by focusing on the organisation of the retinal capillary network itself. Using high-resolution 3 *×* 3-mm imaging and graph-derived metrics, we show that differences in branchpoint density, node number and surviving segment length accompany FAZ deformation in lower-grade disease, even when whole-image vessel length is comparatively preserved. Thus, extending the quantitative density characterisation across Chew grades, the present analysis suggests that network organisation and density capture partly distinct dimensions of the vascular phenotype. This approach may be particularly suited to architectural analysis because the DREAM platform has previously demonstrated detailed depiction of small-calibre vessels and branching patterns, particularly in the DCP, in quantitative cross-device comparisons.[17]

The 3 *×* 3-mm analysis adds layer-specific vessel length, foveal-versus-parafoveal perfusion, FAZ geometry and graph-derived topology using an open-source pipeline. Although the DCP showed the clearest corrected ordinal VLD association, the grade-by-plexus interaction was not significant; layer-specific differences in severity slopes should therefore not be overstated.

Visual acuity provides a supportive clinical context. Acuity worsened steadily with Chew grade, but DCP VLD and parafoveal VAD were not independently associated with logMAR after grade was included in the model. This suggests that, in this modest cross-sectional cohort, vascular and visual abnormalities primarily covaried with the broader multimodal disease state rather than establishing a direct functional relationship with any single OCTA metric. Longitudinal studies incorporating more sensitive functional measures, such as reading speed or microperimetry, will be required to test whether the geometric, topologic, or density phenotypes predict subsequent functional change.

Strengths include prospective recruitment, a contemporaneous healthy cohort, standardised acquisition, transparent OCTAVA processing, independent masked Chew grading, explicit multiplicity control, and mixed-effects models that retain both eyes without treating them as independent. Limitations include the modest sample, sparse grades 1 and 3, cross-sectional design, group differences in axial length, and potential projection or segmentation artefacts in the DCP. All principal models adjusted for axial length, but residual transverse-scaling effects cannot be excluded. OCTAVA-derived topology depends on filtering and skeletonisation settings,[17, 20, 21] and grade 6 includes neovascular disease that can alter flow signals; importantly, the DCP transition remained evident when grade 6 was excluded. The proposed lower-grade-versus-higher-grade framework should therefore be interpreted as a cross-sectional severity pattern, not as proof of a temporal sequence within individual eyes.

In conclusion, high-resolution OCTA across Chew grades supports a multidimensional vascular framework for MacTel severity. Lower grades already show altered FAZ geometry and vascular network topology, whereas higher grades also show regional and whole-image capillary rarefaction. This separation between architectural remodelling and density loss may help explain why a single global OCTA metric incompletely captures MacTel and provides a focused set of candidate phenotypes for longitudinal validation.

## Data availability

Deidentified eye-level data supporting the findings of this study may be made available to researchers for non-commercial scientific use on reasonable request to the corresponding author, subject to institutional approval and a data-use agreement protecting participant confidentiality.

## Supplementary information

Supplementary Tables S1–S6 and Supplementary Figures S1–S2 are provided as a separate Supplementary Information file accompanying this preprint.

## Acknowledgements

All authors are members of the European Reference Network for Rare Eye Diseases (ERN-EYE).

## Author contributions

M.H.: conceptualisation, data curation, formal analysis, methodology, software, visualisation and original draft. D.D.: conceptualisation, data curation and critical revision. S.G.P.: conceptualisation, supervision and critical revision. M.J.G.: conceptualisation, investigation, methodology, project administration, supervision, validation and critical revision. All authors approved the submitted version and accept accountability for the work.

