## Supplementary Information for "Vascular geometry and network topology reveal vascular remodelling beyond density measures across Chew grades in macular telangiectasia type 2"

Supplementary Tables S1–S6 and Supplementary Figures S1–S2.

#### Supplementary Table S1. Key adjusted associations within MacTel

| Analysis | Endpoint | Plexus | Estimate (95% CI) | Adjusted P/q |
|---|---|---|---|---|
| Per Chew grade | Whole-image VLD | SCP | -0.222 (-0.463 to +0.020) | 0.071 |
| Per Chew grade | Whole-image VLD | DCP | -0.352 (-0.583 to -0.121) | 0.007 |
| Per Chew grade | Parafoveal VAD | DCP | -0.593 (-0.943 to -0.242) | 0.007 |
| Per Chew grade | Parafoveal minus foveal VAD | DCP | -0.945 (-1.633 to -0.256) | 0.016 |
| Per Chew grade | FAZ eccentricity | DCP | +0.012 (-0.011 to +0.036) | 0.525 |
| Per Chew grade | FAZ eccentricity | SCP | +0.042 (+0.022 to +0.062) | <0.001 |
| Grades 4–6 minus 0–3 | FD300, percentage points | DCP | -2.545 (-4.478 to -0.611) | 0.050 |
| Grades 4–6 minus 0–3 | FD300, percentage points | SCP | -2.657 (-4.621 to -0.693) | 0.050 |

Models adjusted for age, sex and axial length and included a participant-level random intercept. VLD, vessel length density; VAD, vessel area density; FAZ, foveal avascular zone; FD300, vessel density within 300 $\mu$m of the FAZ.

### Supplementary Table S2. Network topology differences from healthy controls

| Severity | Endpoint | Plexus | Adjusted difference (95% CI) | BH q |
|---|---|---|---|---|
| Early (0–2) | Branchpoint density | DCP | -0.246 (-0.606 to +0.113) | 0.211 |
| Early (0–2) | Branchpoint density | SCP | -0.446 (-0.792 to -0.100) | 0.022 |
| Early (0–2) | Mean segment length | DCP | +0.535 (-0.240 to +1.309) | 0.211 |
| Early (0–2) | Mean segment length | SCP | +1.330 (+0.149 to +2.510) | 0.042 |
| Early (0–2) | Node count | DCP | -64.980 (-250.537 to +120.577) | 0.516 |
| Early (0–2) | Node count | SCP | -182.679 (-352.558 to -12.800) | 0.049 |
| Intermediate (3–4) | Branchpoint density | DCP | -0.237 (-0.744 to +0.270) | 0.400 |
| Intermediate (3–4) | Branchpoint density | SCP | -0.688 (-1.136 to -0.240) | 0.008 |
| Intermediate (3–4) | Mean segment length | DCP | +0.351 (-0.777 to +1.479) | 0.538 |
| Intermediate (3–4) | Mean segment length | SCP | +2.255 (+0.682 to +3.828) | 0.011 |
| Intermediate (3–4) | Node count | DCP | -270.504 (-500.994 to -40.013) | 0.036 |
| Intermediate (3–4) | Node count | SCP | -319.862 (-542.531 to -97.193) | 0.011 |
| Advanced (5–6) | Branchpoint density | DCP | -0.762 (-1.167 to -0.357) | 0.003 |
| Advanced (5–6) | Branchpoint density | SCP | -0.615 (-0.991 to -0.239) | 0.005 |
| Advanced (5–6) | Mean segment length | DCP | +1.544 (+0.658 to +2.430) | 0.005 |
| Advanced (5–6) | Mean segment length | SCP | +2.429 (+1.131 to +3.727) | 0.003 |
| Advanced (5–6) | Node count | DCP | -322.063 (-520.231 to -123.895) | 0.005 |
| Advanced (5–6) | Node count | SCP | -303.320 (-488.819 to -117.822) | 0.005 |

### Supplementary Table S3. FAZ geometry differences from healthy controls

| Severity | Endpoint | Plexus | Adjusted difference (95% CI) | BH q |
|---|---|---|---|---|
| Advanced (5–6) | FAZ acircularity index | DCP | +0.137 (+0.073 to +0.201) | <0.001 |
| Advanced (5–6) | FAZ acircularity index | SCP | +0.227 (+0.171 to +0.284) | <0.001 |
| Early (0–2) | FAZ acircularity index | DCP | +0.073 (+0.018 to +0.127) | 0.034 |
| Early (0–2) | FAZ acircularity index | SCP | +0.132 (+0.083 to +0.181) | <0.001 |
| Intermediate (3–4) | FAZ acircularity index | DCP | +0.107 (+0.023 to +0.191) | 0.045 |
| Intermediate (3–4) | FAZ acircularity index | SCP | +0.134 (+0.063 to +0.206) | 0.002 |
| Advanced (5–6) | FAZ eccentricity | DCP | +0.108 (+0.022 to +0.195) | 0.047 |
| Advanced (5–6) | FAZ eccentricity | SCP | +0.242 (+0.160 to +0.324) | <0.001 |
| Early (0–2) | FAZ eccentricity | DCP | +0.064 (-0.009 to +0.138) | 0.222 |
| Early (0–2) | FAZ eccentricity | SCP | +0.048 (-0.023 to +0.119) | 0.349 |
| Intermediate (3–4) | FAZ eccentricity | DCP | +0.074 (-0.040 to +0.188) | 0.375 |
| Intermediate (3–4) | FAZ eccentricity | SCP | +0.161 (+0.056 to +0.266) | 0.014 |

#### Supplementary Table S4. Regional VAD associations with Chew severity

| Analysis | Region | Plexus | Estimate (95% CI) | BH q |
|---|---|---|---|---|
| Ordinal slope | Foveal circle | DCP | +0.228 (-0.506 to +0.962) | 0.715 |
| Ordinal slope | Inferior hemi-ring | DCP | -0.715 (-1.133 to -0.298) | 0.007 |
| Ordinal slope | Superior hemi-ring | DCP | -0.424 (-0.878 to +0.030) | 0.152 |
| Ordinal slope | Inferior quadrant | DCP | -0.011 (-0.754 to +0.733) | 0.977 |
| Ordinal slope | Nasal quadrant | DCP | -0.813 (-1.233 to -0.394) | 0.005 |
| Ordinal slope | Parafoveal rim | DCP | -0.593 (-0.943 to -0.242) | 0.007 |
| Ordinal slope | Superior quadrant | DCP | -0.218 (-0.643 to +0.208) | 0.495 |
| Ordinal slope | Temporal quadrant | DCP | -0.584 (-1.031 to -0.137) | 0.037 |
| Ordinal slope | Foveal circle | SCP | -0.060 (-0.844 to +0.724) | 0.937 |
| Ordinal slope | Inferior hemi-ring | SCP | -0.274 (-0.675 to +0.127) | 0.311 |
| Ordinal slope | Superior hemi-ring | SCP | -0.382 (-0.854 to +0.089) | 0.220 |
| Ordinal slope | Inferior quadrant | SCP | -0.213 (-1.021 to +0.595) | 0.737 |
| Ordinal slope | Nasal quadrant | SCP | -0.674 (-1.141 to -0.206) | 0.022 |
| Ordinal slope | Parafoveal rim | SCP | -0.368 (-0.715 to -0.020) | 0.103 |
| Ordinal slope | Superior quadrant | SCP | -0.080 (-0.505 to +0.344) | 0.805 |
| Ordinal slope | Temporal quadrant | SCP | -0.180 (-0.572 to +0.211) | 0.520 |
| Grades 4–6 minus 0–3 | Inferior hemi-ring | DCP | -2.655 (-4.333 to -0.977) | 0.016 |
| Grades 4–6 minus 0–3 | Nasal quadrant | DCP | -3.462 (-5.143 to -1.782) | 0.002 |
| Grades 4–6 minus 0–3 | Parafoveal rim | DCP | -2.190 (-3.597 to -0.782) | 0.016 |
| Grades 4–6 minus 0–3 | Nasal quadrant | SCP | -2.642 (-4.489 to -0.795) | 0.024 |

The table displays all ordinal slopes and the transition contrasts that remained significant after multiplicity adjustment. Full non-significant transition contrasts are retained in the statistical output workbook used for numerical verification.

#### Supplementary Table S5. FAZ and FD300 associations within MacTel

| Analysis | Endpoint | Plexus | Estimate (95% CI) | BH q |
|---|---|---|---|---|
| Ordinal slope | FAZ acircularity index | DCP | +0.016 (-0.005 to +0.036) | 0.282 |
| Ordinal slope | FAZ area | DCP | -0.009 (-0.031 to +0.014) | 0.573 |
| Ordinal slope | FAZ eccentricity | DCP | +0.012 (-0.011 to +0.036) | 0.525 |
| Ordinal slope | FD300 | DCP | -0.436 (-0.941 to +0.069) | 0.253 |
| Ordinal slope | FAZ acircularity index | SCP | +0.016 (-0.002 to +0.034) | 0.253 |
| Ordinal slope | FAZ area | SCP | +0.002 (-0.020 to +0.025) | 0.842 |
| Ordinal slope | FAZ eccentricity | SCP | +0.042 (+0.022 to +0.062) | <0.001 |
| Ordinal slope | FD300 | SCP | -0.603 (-1.109 to -0.098) | 0.092 |
| Grades 4–6 minus 0–3 | FAZ acircularity index | DCP | +0.060 (-0.020 to +0.139) | 0.274 |
| Grades 4–6 minus 0–3 | FAZ area | DCP | -0.049 (-0.137 to +0.040) | 0.475 |
| Grades 4–6 minus 0–3 | FAZ eccentricity | DCP | +0.048 (-0.042 to +0.139) | 0.475 |
| Grades 4–6 minus 0–3 | FD300 | DCP | -2.545 (-4.478 to -0.611) | 0.050 |
| Grades 4–6 minus 0–3 | FAZ acircularity index | SCP | +0.058 (-0.014 to +0.130) | 0.248 |
| Grades 4–6 minus 0–3 | FAZ area | SCP | -0.023 (-0.113 to +0.067) | 0.687 |
| Grades 4–6 minus 0–3 | FAZ eccentricity | SCP | +0.160 (+0.084 to +0.236) | 0.001 |
| Grades 4–6 minus 0–3 | FD300 | SCP | -2.657 (-4.621 to -0.693) | 0.050 |

### Supplementary Table S6. Sensitivity and visual-function analyses

| Analysis | Endpoint | Estimate (95% CI) | P |
|---|---|---|---|
| Exclude grade 6 | SCP VLD transition | -0.801 (-1.797 to +0.195) | 0.112 |
| Exclude grade 6 | DCP VLD transition | -1.171 (-2.177 to -0.165) | 0.024 |
| Add logMAR visual acuity | DCP VLD slope per grade | -0.339 (-0.596 to -0.083) | 0.010 |

| Model | Predictor | Estimate (95% CI) | Raw P | Holm P |
|---|---|---|---|---|
| Clinical severity | Chew grade | +0.071 (+0.036 to +0.107) | <0.001 | — |
| Without Chew grade | DCP whole-image VLD | -0.066 (-0.155 to +0.023) | 0.141 | 0.141 |
| With ordinal Chew grade | DCP whole-image VLD | -0.014 (-0.101 to +0.074) | 0.755 | 0.797 |
| Without Chew grade | DCP parafoveal VAD | -0.092 (-0.176 to -0.007) | 0.034 | 0.068 |
| With ordinal Chew grade | DCP parafoveal VAD | -0.037 (-0.123 to +0.050) | 0.398 | 0.797 |

### Supplementary Figure S1. Multi-domain structural transition across the grade 3-4 boundary

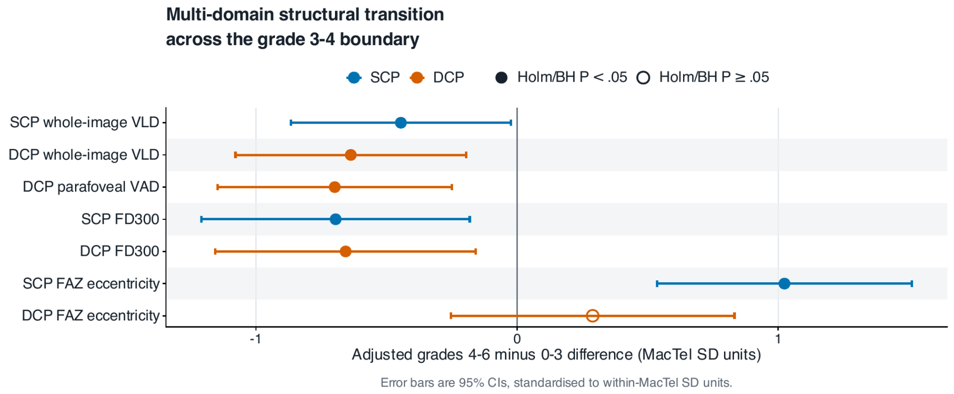


Adjusted grades 4–6 minus 0–3 differences are standardised to within-MacTel SD units. Filled symbols indicate Holm/BH P<0.05; open symbols indicate P≥0.05. Error bars are 95% confidence intervals.

#### Supplementary Figure S2. Regional vessel area density by quadrant and hemifield

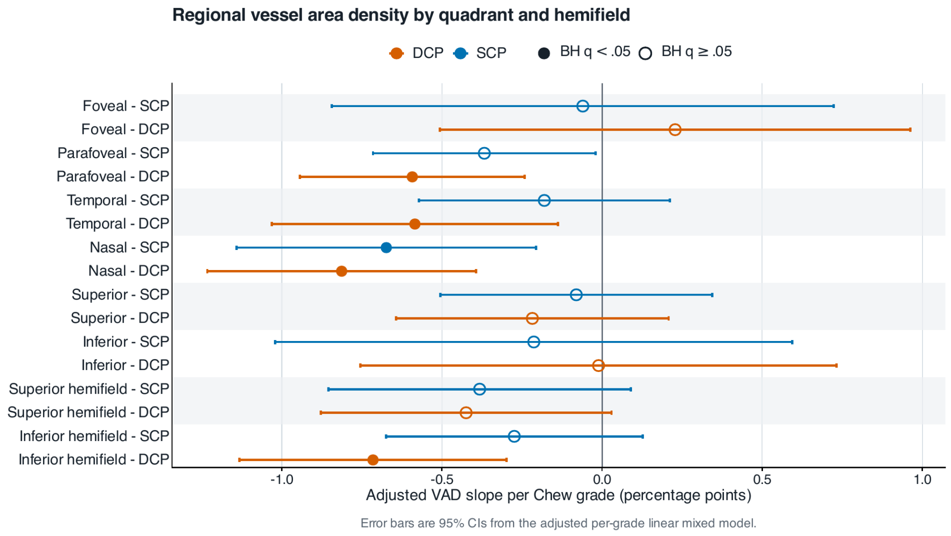


Adjusted VAD slope per Chew grade in percentage points. Filled symbols indicate BH $q<0.05$; open symbols indicate $q\geq0.05$. Error bars are 95% confidence intervals from the adjusted per-grade mixed model.
